# ^124^I Quantification with LAFOV PET: Calibration, Bias and Systematic Performance Evaluation

**DOI:** 10.64898/2026.07.28.26359126

**Authors:** Elisa Körner, Walter Jentzen, Pia M. Linder, Jorge Cabello, Johannes Schwenck, Ivo Rausch, Christian la Fougère, Fabian P. Schmidt

## Abstract

**Background:** Quantitative ¹²⁴I PET imaging is challenged by low positron branching ratio, prompt gamma emissions, and limited count statistics. Long axial field-of-view (LAFOV) PET systems provide substantially increased sensitivity, potentially enabling more robust imaging in terms of quantitative accuracy and noise mitigation under these conditions. This study aimed to systematically evaluate image quality, quantitative accuracy, and sources of bias in low-count ¹²⁴I PET for varying acquisition times across the preparation and imaging pipeline, with particular focus on scatter/prompt gamma correction and dose calibrator calibration.

**Methods:** Multiple phantoms were employed in the current study, namely the NEMA IQ phantom (sphere-to-background ratio 20:1) and cylindrical phantoms of different sizes representing different scatter geometries. All phantoms were filled with low activity concentrations typical of clinical imaging (about 0.4 kBq/mL background, corresponding to a 37 MBq ^124^I administration in a 70 kg patient imaged 24 h post-injection). Contrast recovery, recovery coefficients, image noise as coefficient of variation (CV), and lung residual error were assessed. Data were acquired on a LAFOV PET/CT scanner (Biograph Vision Quadra, Siemens Healthineers) and reconstructed using single scatter simulation with tail fitting (SSS-TF) and an alternative maximum-likelihood scatter scaling approach (SSS-MLSS). The impacts of acquisition time (15 min vs. 30 min) and object size on image quality and quantification were evaluated. Dose calibrator performance and inter-device consistency were assessed across 0.5–60 MBq range of activity. A representative ^124^I PET scan of a patient with metastatic differentiated thyroid cancer (DTC) was included to assess lesion detectability and quantification at reduced scan durations.

**Results:** Image quality remained robust under low-count ^124^I conditions (0.4 kBq/mL) with a CV of 15.8% at 30 min, which is consistent with EANM/EARL recommendations and comparable to matched low-count ¹⁸F acquisitions (15.4% at 30 min). Reducing acquisition time to 15 min increased noise but preserved contrast and recovery (≤2.1% and <±2%). Using SSS-TF, activity concentration was underestimated for ^124^I, particularly in the background of the NEMA IQ phantom (83.0% for ¹²⁴I vs. 102.7% for ¹⁸F). SSS-MLSS improved background recovery (95.8%) while maintaining sphere recovery, yielding more consistent quantification. Size-dependent effects were observed, with underestimation in larger objects (86.1% phantom ø=8 cm vs. 81.4% phantom ø=20 cm using SSS-TF), which was reduced using SSS-MLSS (87.0% vs. 96.1%, respectively). Dose calibrator measurements showed high stability and low inter-device variability (≤2.3%). In the patient dataset, lesion detectability and quantification remained stable across reconstruction methods and scan durations down to 5 min.

**Conclusion:** LAFOV PET enables robust low-count ¹²⁴I imaging with preserved image quality and quantification, allowing the reduction of acquisition times to ≤15 min. Quantitative accuracy is primarily impaired by scatter including prompt gamma coincidence correction and object geometry, while calibration-related effects are minor under controlled conditions.

## Introduction

^124^I is a positron-emitting radionuclide with established applications in immuno-PET [1,2] and theragnostic workflows, particularly for pre-therapeutic assessment and individualized dosimetry in ^131^I-based therapies. Clinically, ^124^I PET is used to guide ^131^I meta-Iodobenzylguanidine (miBG) therapy in neural crest-derived tumors [3,4] and to optimize ^131^I therapy in differentiated thyroid cancer (DTC) [5,6]. Compared with conventional gamma camera imaging using ^123^I [4] or ^131^I [7,8], ^124^I PET provides superior sensitivity and lesion detectability.

However, quantitative ¹²⁴I PET imaging remains technically challenging due to its low positron branching ratio of 22.5% and the need for relatively low administered activities in DTC (about 37 MBq [9]) to avoid stunning effects, resulting in limited count statistics.

Improving PET system sensitivity is therefore essential to compensate for the limited count statistics in ¹²⁴I imaging. The transition from analog to digital silicon photomultiplier (SiPM)-based PET/CT systems, has already led to substantial improvements in lesion detectability and reduced acquisition times [9,10]. In addition, the time-of-flight (TOF) performance of modern PET systems further enhances the signal-to-noise ratio by increasing the effective sensitivity [11].

Building on these developments, long axial field-of-view (LAFOV) PET systems extend the same detector technology to a substantially larger axial coverage, resulting in a further increase in sensitivity. For example, the Biograph Vision Quadra (Siemens Healthineers, Knoxville, TN, USA) achieves an approximately tenfold higher sensitivity than the Biograph Vision 600, as measured per the NEMA standard (176 cps/kBq [12] vs. 16.4 cps/kBq [13]). This increase in sensitivity is expected to further improve the intrinsic limitations of ¹²⁴I, to enhance image quality and to produce more robust quantification, particularly for reduced acquisition times and low administered activities.

Beyond conventional imaging, the prompt gamma emission of ¹²⁴I enables applications such as positronium life time imaging [14,15] and tracer multiplexing for simultaneous dual-tracer PET acquisitions [16]. However, despite these developments, a systematic evaluation of quantitative ¹²⁴I imaging on LAFOV PET systems is currently lacking. In particular, the impact of the entire quantification chain, including activity measurement in the dose calibrator, cross-calibration using ^18^F, acquisition parameters, and data corrections in the image reconstruction, has not been comprehensively assessed for this non-standard radionuclide.

Importantly, results from low-count ¹⁸F imaging studies [17,18] cannot be directly transferred to ¹²⁴I due to its fundamentally different decay characteristics. In contrast to ¹⁸F, accurate ¹²⁴I quantification requires dedicated correction for the prompt gamma coincidence contamination [19–21], and is further affected by the increased positron range [22], leading to reduced spatial resolution and recovery coefficient in small objects. In DTC, ¹²⁴I imaging often exhibits highly specific focal tracer uptake with relatively low background activity [9], resulting in imaging conditions that differ substantially from standard applications, such as [¹⁸F]FDG PET.

Therefore, we performed dedicated phantom experiments designed to mimic ¹²⁴I patient scan conditions on a LAFOV PET/CT system. In this study, we systematically evaluated quantitative accuracy, recovery characteristics, and image quality across varying acquisition durations, with a particular focus on short scan protocols, and compared these results to corresponding low-count ¹⁸F reference measurements (corrected for positron branching ratio).

As a prerequisite for accurate quantification, we further assessed factors affecting the complete preparation and imaging chain, including activity measurement accuracy in the dose calibrator (calibration accuracy, activity dependence, and inter-device variability), ^18^F cross-calibration with the PET system, and image reconstruction-specific corrections. A particular focus was on scatter correction, which also intrinsically corrects for prompt gamma coincidences, by comparing the clinically established single-scatter simulation with tail fitting (SSS-TF) to the investigative maximum-likelihood scatter scaling approach (SSS-MLSS) [23,24]. By systematically examining these factors, we aimed to identify potential sources of bias in ¹²⁴I quantification.

We investigated whether the increased sensitivity of LAFOV PET enables accurate and robust quantification of ¹²⁴I under clinically realistic and reduced acquisition conditions. Finally, we demonstrated on a clinical case the potential of shortened acquisition times and assessed the impact of reconstruction choices on image quality and quantification.

## Materials and Methods

### Study Design - Assessment of Factors affecting Quantification Estimation

Quantitative PET imaging is influenced by multiple factors across the entire imaging chain, ranging from activity preparation to image reconstruction. In this study, we systematically assessed these factors as illustrated in Figure 1, distinguishing between the contributions related to the dose calibrator and those related to the scanner system.

**Figure 1.**
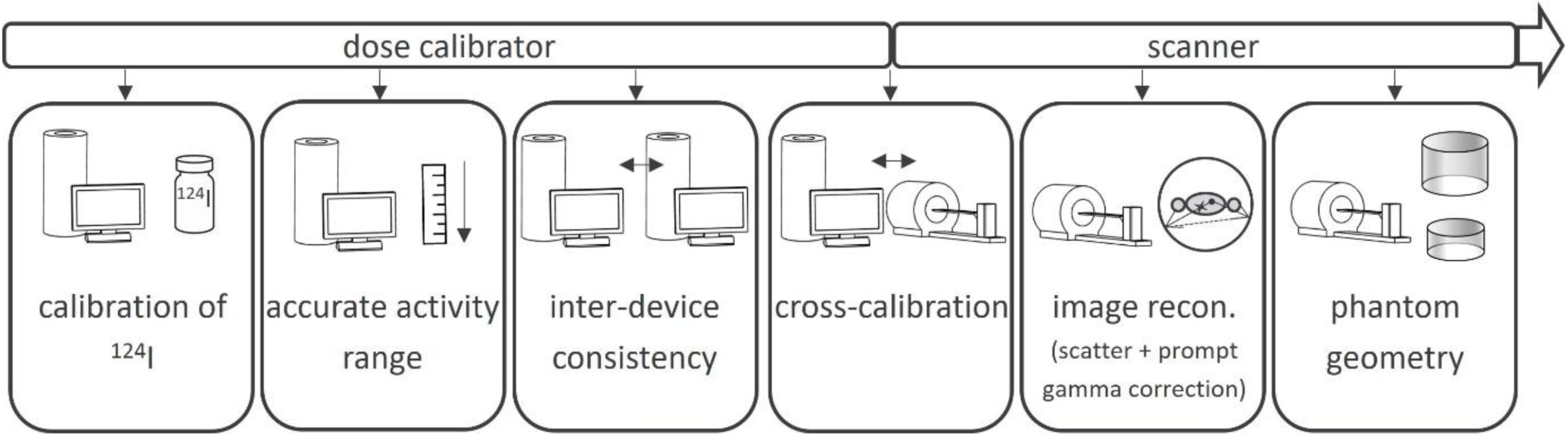
Overview of factors affecting quantitative accuracy assessed in this study, structured along the imaging chain from activity preparation (dose calibrator) to PET acquisition and image reconstruction.

On the dose calibrator side, this includes calibration accuracy, activity dependences, and inter-device variabilities. On the scanner side, factors such as ^18^F cross-calibration, image reconstruction-specific corrections, and phantom geometry are considered. This structured approach enabled a comprehensive evaluation of potential sources of bias in ¹²⁴I quantification.

#### Calibration of ^124^I

Accurate measurement of ^124^I in dose calibrators is essential, as the measured activity depends on vial geometry, material and position in the dose calibrator [25]. In this study, ^124^I activity was calibrated using a 2 mL ^124^I solution in a P6 vial (Amersham International, Buckinghamshire, UK) with a CRC-15R dose calibrator (Capintec Inc., Ramsey, NJ, USA). The calibration was performed following the protocol described in literature [26] using the identical dose calibrator, vial geometry, measurement position, and activity range. Under these conditions, the authors reported deviations of <±3% between CRC-15R measurements and reference values traceable to the Physikalisch-Technische Bundesanstalt (PTB, national authority for radiation measurement standards).

To ensure reproducibility, each activity measurement was performed in three independent repetitions, with the vial removed and repositioned in the dose calibrator between measurements, and the mean value was used for calibration. Two additional dose calibrators (both VDC-405, V3.30, Comecer S.p.A., Castel Bolognese, Italy), referred to as VDC-A and VDC-B, were subsequently calibrated against the CRC-15R using identical vial geometries.

#### Dose Calibrator Activity-Level Accuracy and Inter-Device Variability

The activity-level accuracy (linearity) of the dose calibrators was evaluated over a clinically relevant activity range of 0.5-60 MBq for the 2 mL ¹²⁴I solution. Repeated measurements were performed daily over a period of one month and compared to the decay-corrected initial activity.

Inter-device consistency was assessed by comparing measurements obtained from VDC-A and VDC-B to those of the CRC-15R. This allowed evaluation of agreement across different calibrator models and manufacturers.

#### Dose Calibrator Cross-calibration with the PET Scanner

Quantitative PET imaging requires accurate cross-calibration between the dose calibrator and the PET/CT system. In this study, the PET/CT system (Biograph Vision Quadra, Siemens Healthineers, Knoxville, TN, USA) was cross-calibrated to the dose calibrator according to EARL standards (European Association of Nuclear Medicine Research Ltd.) [27] using a 20-cm-diameter cylindrical phantom containing approximately 70 MBq of ^18^F. All dose calibrators employed in this study (VDC-405, CRC-15R) were verified against a reference calibration standard and cross-calibrated.

### Phantom Experiments and Acquisition Protocol

Image quality, noise, and quantitative accuracy for ^124^I were assessed using a NEMA IQ phantom. Phantom preparation deviated from the NEMA NU-2-2018 protocol to reflect clinically realistic ^124^I imaging conditions (Table 1).

**Table 1.**
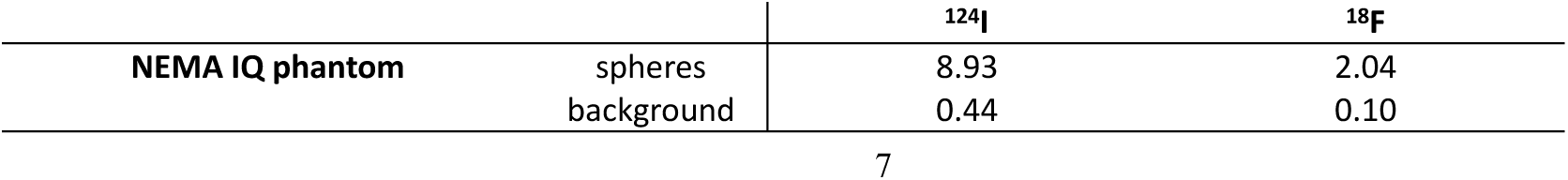
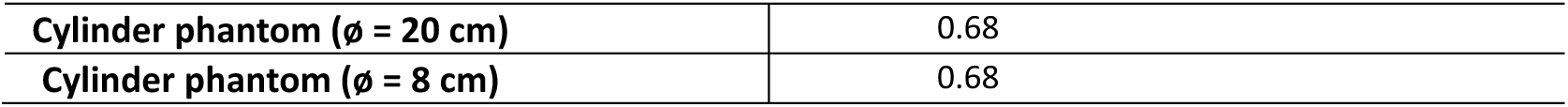
Prepared activity concentrations [kBq/mL] for the NEMA IQ phantom spheres and background compartment scanned with ^124^I and ^18^F and the ^124^I cylinder phantoms of two different sizes.

| | | $^{124}\text{I}$ | $^{18}\text{F}$ |
| --- | --- | --- | --- |
| NEMA IQ phantom | spheres | 8.93 | 2.04 |
|  | background | 0.44 | 0.10 |

|  |  |
| --- | --- |
| Cylinder phantom ($\phi$ = 20 cm) | 0.68 |
| Cylinder phantom ($\phi$ = 8 cm) | 0.68 |

A higher sphere-to-background ratio (SBR) of 20.2:1 was selected to mimic focal uptake observed in DTC [9]. In addition, a substantially lower background activity concentration of 0.44 kBq/mL was used, compared with 5.30 kBq/mL for ^18^F in NEMA NU-2-2018, corresponding to a typical 37 MBq ^124^I administration in a 70 kg patient imaged 24 h post-injection. The phantom comprised six fillable spheres (diameters 37, 28, 22, 17, 13, and 10 mm) with an activity concentration of 8.93 kBq/mL. The preparation included filling the phantom with purified water containing non-radioactive iodine to saturate possible binding sites and prevent ^124^I adhesion to the phantom walls.

For comparison, a corresponding measurement was performed using ¹⁸F under matched high-SBR and low-count conditions. Activity concentrations were adjusted to 0.10 kBq/mL in the background and 2.04 kBq/mL in the spheres to achieve comparable count statistics, accounting for differences in positron branching ratios (^124^I: 22.5% vs. ^18^F: 96.7%).

A 30 min acquisition was performed for both phantoms (one filled with ^124^I and the other one with ^18^F), with the spheres centered in the axial field of view (FOV) and the long axis of the lung insert aligned with the long axis of the PET detectors and centered in the transaxial FOV. To explore size-dependent effects on scatter correction and quantification, additional measurements were performed using cylindrical phantoms of two different sizes: ø=20 cm, length= 20 cm and ø=8 cm, length=16 cm. ¹²⁴I activity concentrations were selected to be in a comparable range to the NEMA IQ phantom experiments (around 0.68 kBq/mL in both cylindrical phantoms) (Table 1). The identical ¹²⁴I solution was used for both cylindrical phantoms to minimize potential confounding effects on absolute quantification.

Both phantoms were positioned at the center of the axial and transaxial FOV with the long axis aligned with the PET system axis, and PET data were acquired for 30 min.

### Image Reconstruction

All datasets were reconstructed using a vendor-provided software (e7 tools with version VR20, Siemens Healthineers, Knoxville, TN, USA) following a reconstruction protocol routinely employed for clinical ¹²⁴I imaging at our institution. An ordinary Poisson ordered-subsets expectation-maximization algorithm with four iterations and five subsets was employed, including point-spread-function modelling and time-of-flight information. Reconstructions were performed in ultra-high sensitivity mode using a 440×440×645 matrix with a voxel size of 1.65×1.65×1.65 mm³, without post-filtering.

CT-based (120 kVp, 50 mAs) attenuation correction, prompt gamma correction and scatter correction using 3D SSS-TF were applied as implemented in the clinical reconstruction. In addition, an alternative scatter correction approach based on maximum-likelihood scatter scaling (SSS-MLSS) was applied to evaluate its impact on quantification. In the employed reconstruction framework, prompt gamma correction is intrinsically linked to the scatter correction scaling procedure.

In SSS-TF, the simulated scatter distribution is scaled using tail fitting in sinogram regions identified outside the attenuation map, assumed to contain predominantly scattered events [28]. In contrast, SSS-MLSS, originally proposed by Panin et al. [23] and further refined by Rezaei et al. [24], updates the scatter scaling factors iteratively using a maximum-likelihood approach based on the full emission sinogram, thereby incorporating information from the entire dataset. In addition, the list mode data of the ^124^I NEMA IQ phantom scan was rebinned to simulate a shorter scan time of 15 min.

### Quantitative Image Analysis

Image quality was evaluated by contrast recovery, derived from spherical volumes of interest (VOIs) corresponding to the phantom sphere diameters (10-37 mm) and a boxed-shaped 150×15×160 mm³ VOI placed in the background compartment below the spheres. Contrast recovery coefficients (CRCs) were calculated as the measured SBR, normalized to the prepared SBR.

Image noise was quantified using the coefficient of variation (CV), defined as the ratio of the voxel-wise standard deviation to the measured mean activity concentration within the background VOI.

To assess potential limitations of scatter correction, residual activity in the lung insert was evaluated. A cylindrical VOI (30 mm diameter, 160 mm length) was placed within the lung insert (diameter ∼45 mm), and the lung residual error (LRE) was determined as the ratio of its mean activity concentration to the mean activity concentration of the background VOI.

Quantitative accuracy was evaluated using absolute recovery coefficients (RCs) for both the spheres and the background compartment, defined as the ratio of measured to prepared activity concentration.

For the cylindrical phantom experiments, RCs were calculated within central VOIs (phantom with ø = 20 cm, VOI with ø = 16 cm and phantom with ø = 8 cm, VOI with ø = 5 cm) to assess size-dependent effects on quantification.

All delineation and analysis were performed using AMIDE software version 1.04 [29].

### Patient Case

To illustrate the clinical relevance of the phantom findings, a representative patient with metastatic DTC was included. PET/CT imaging was performed on a Siemens Biograph Vision Quadra approximately 24 h after oral administration of 37 MBq ¹²⁴I. A diagnostic CT (120 kVp, 369 mAs) was followed by a 15 min PET acquisition.

To assess the impact of acquisition time and reconstruction on lesion detectability and quantification, list mode data were reconstructed analogously to the phantom study with SSS-TF and SS-MLSS and rebinned to simulate shorter scan durations of 10 min and 5 min.

Multiple iodine-avid bone metastases (iliac bone, humerus, vertebral bodies C1, C4, T9, and L1) were evaluated. Lesion uptake was quantified using SUV_mean_ and SUV_max_ derived from semi-automated VOIs (50% isocontour threshold) in Affinity Viewer (Version 2.18, Hermes Medical Solutions, Sweden).

## Results

### Activity-Level Accuracy and Inter-Device Variability of Dose Calibrators

The measured activities of all dose calibrators showed good agreement (linearity) with the decay-corrected reference values down to 1 MBq ^124^I (Figure 2). Between 1 MBq and 60 MBq, percentage mean deviations between measured and decay-corrected activities were 0.8 ± 1.2% (CRC-15R), 0.6 ± 0.6% (VDC-A) and 1.0 ± 0.7% (VDC-B). Corresponding maximum percentage deviations were 5.7% (1.70 vs. 1.61 MBq), 2.7% (1.02 vs. 0.99 MBq) and -2.9% (1.31 vs. 1.35 MBq).

**Figure 2.**
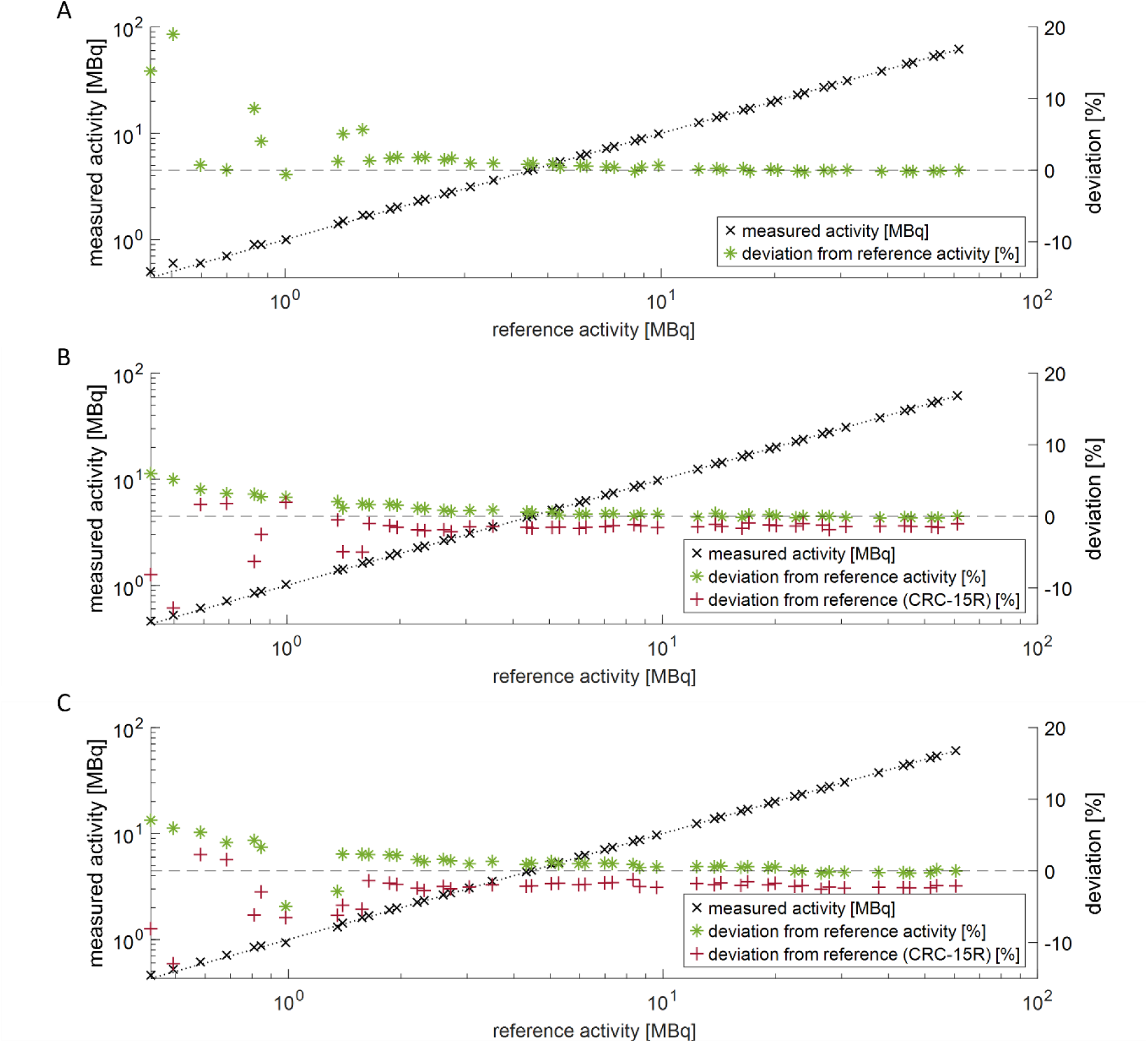
Measured activity as a function of reference activity (calculated via decay from the initial 60 MBq activity) for the CRC-15R (A), VDC-A (B), and VDC-B (C). Black markers indicate measured activity, while green markers show the relative deviation from the reference activity. Red markers (B–C) represent deviations relative to the CRC-15R as the reference. Measurements were performed over an activity range of approximately 0.5–60 MBq.

The inter-device comparisons relative to the CRC-15R demonstrated good consistency across calibrators with mean inter-device deviations of 1.6 ± 0.8% (VDC-A) and 2.3 ± 1.0% (VDC-B), and maximum deviations of -5.0% (1.62 MBq vs. 1.70 MBq) and -6.2% (1.31 vs. 1.40 MBq) for activities > 1MBq.

Overall, these results indicate stable and consistent activity measurements between devices down to 1 MBq, supporting reliable activity quantification for subsequent PET measurements.

### Image Quality, Noise and Contrast Recovery

Image quality of the 30 min ¹⁸F NEMA IQ phantom scan (Figure 3G) was comparable to the ¹²⁴I acquisition reconstructed with SSS-TF (Figure 3A), which was also reflected by similar image noise levels, with CVs of 15.4% for ¹⁸F and 15.8% for ¹²⁴I.

**Figure 3.**
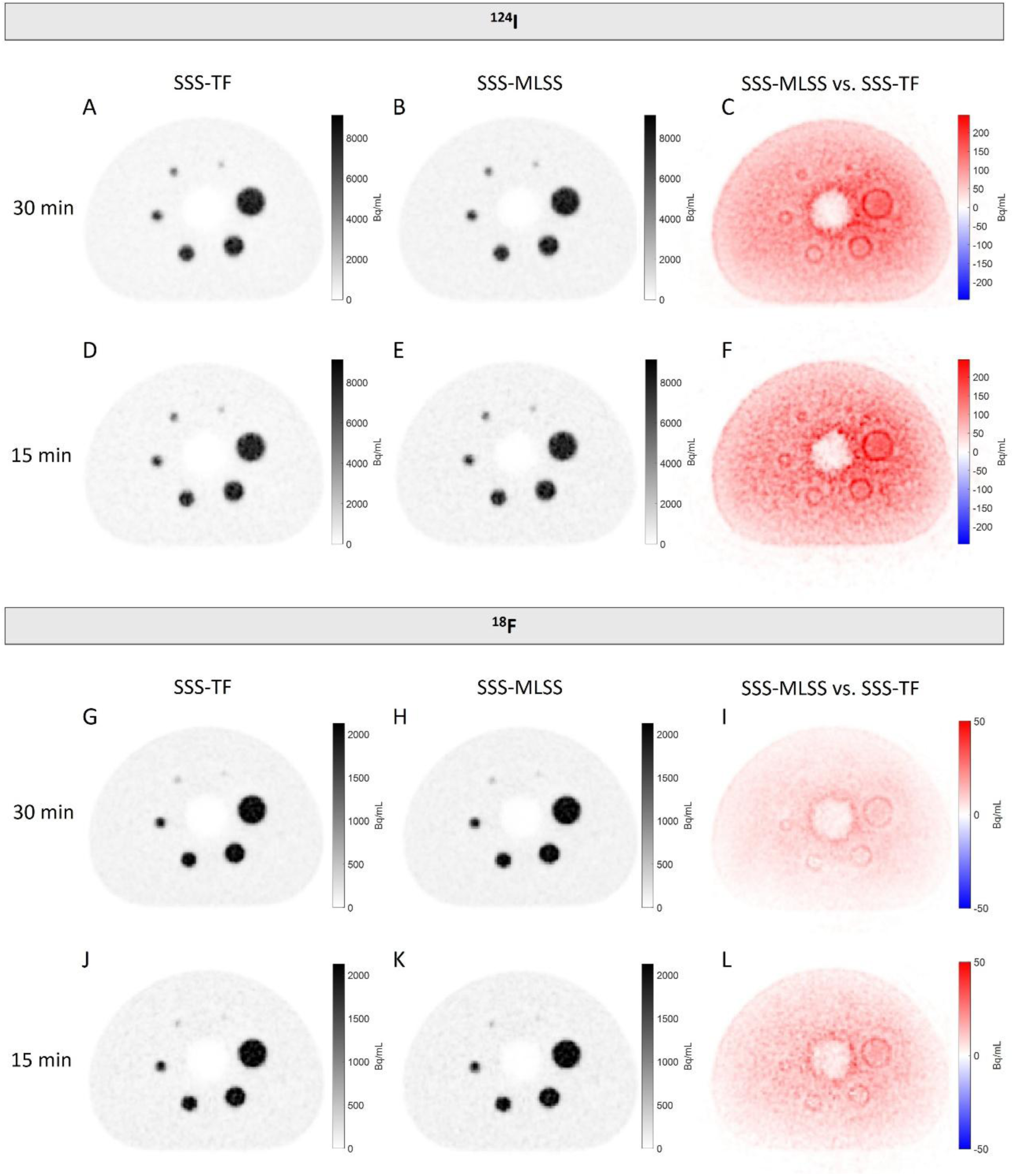
Representative transverse slices of the NEMA IQ phantom. (A–C) ¹²⁴I (30 min) reconstructed with SSS-TF (A), SSS-MLSS (B), and corresponding difference image (SSS-MLSS minus SSS-TF) (C). (D–F) ¹²⁴I (15 min) reconstructed with SSS-TF (D), SSS-MLSS (E), and corresponding difference image (F). (G–I) ^18^F (30 min) reconstructed with SSS-TF (G), SSS-MLSS (H), and corresponding difference image (I). (J–L) ^18^F (15 min) reconstructed with SSS-TF (J), SSS-MLSS (K), and corresponding difference image (L).

The noise characteristics observed in SSS-MLSS reconstructions (Figure 3B, CV: 15.4% for ^124^I) were comparable to SSS-TF, indicating no relevant difference in image noise between the scatter correction methods.

Reduction of scan time to 15 min resulted in increased image noise (CV: 21.9% for SSS-TF and 21.6% for SSS-MLSS; Figure 3D/E). Despite this increase, overall image quality remained high, and all spheres, including the smallest (10 mm), remained clearly identifiable (Figure 3D/E). The difference images between SSS-MLSS and SSS-TF reconstructions (Figure 3C, F) revealed a systematic increase in the reconstructed ^124^I activity for SSS-MLSS across the entire phantom. This increase was particularly pronounced at the edges of the spheres, indicating differences in scatter estimation which affect local contrast.

In addition, elevated activity was observed within the lung insert for SSS-MLSS, consistent with an increased lung residual error of 7.0% compared to 1.9% for SSS-TF (^18^F: 6.3% using SSS-MLSS and 4.1% using SSS-TF).

In contrast to the pronounced differences observed for ¹²⁴I, the corresponding ¹⁸F difference images (Figure 3I,L) showed only minor deviations between SSS-MLSS and SSS-TF. This finding suggests that the observed discrepancies in ¹²⁴I are primarily related to prompt gamma correction rather than to scatter correction itself.

Consistent with these observations, contrast recovery of ^124^I remained stable under scan time reduction, with deviations of at most -2.1% for the 10 mm sphere between 30 min and 15 min CRCs using SSS-TF (Figure 4).

**Figure 4.**
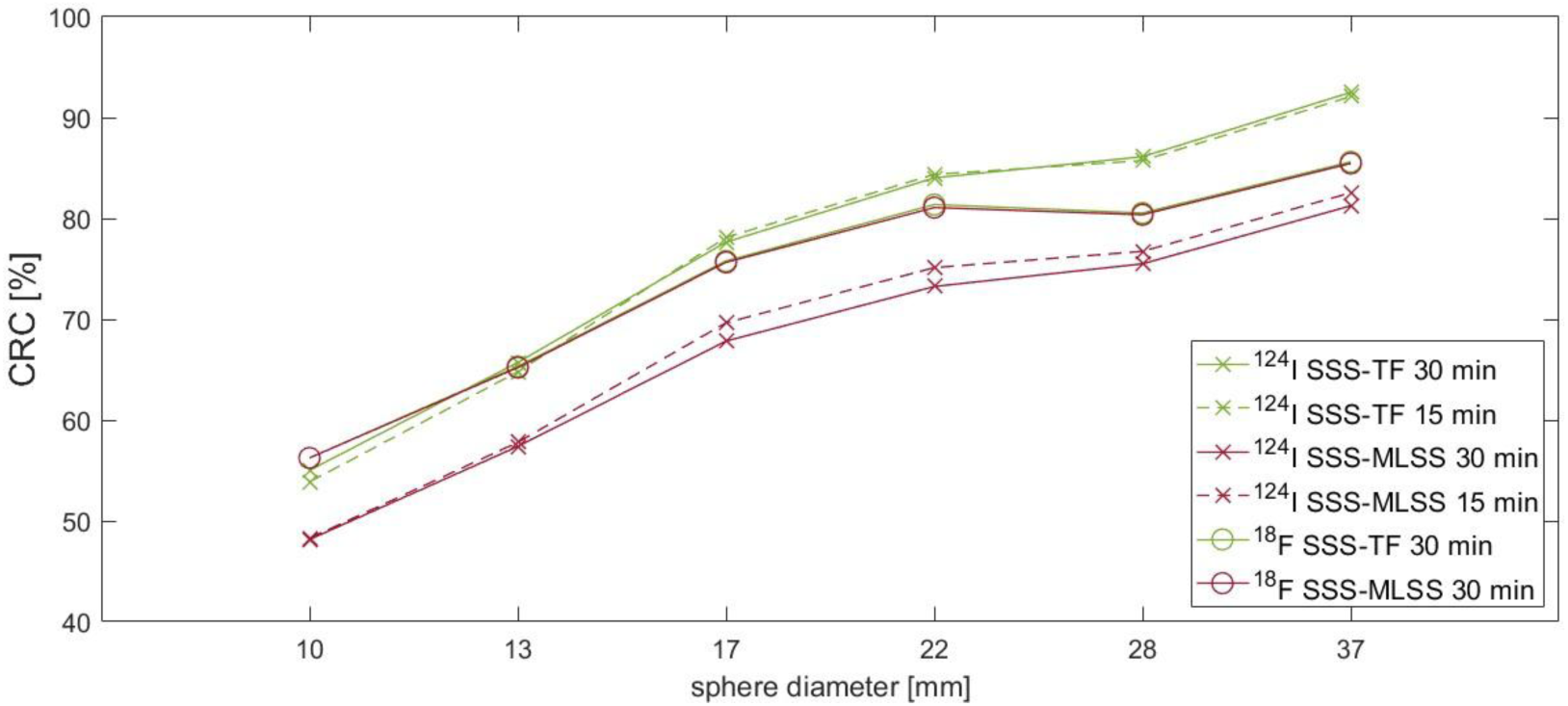
Contrast recovery coefficients (CRC, [%]) as a function of sphere diameter (10-37 mm) for the NEMA IQ phantom. CRCs for ^124^I (30 min and 15 min) reconstructed using SSS-TF and SSS-MLSS for scatter correction, with CRCs for ^18^F (30 min, SSS-TF and SSS-TF) included for comparison.

Representative CRC values for ^124^I using SSS-TF were 86.1% (28 mm), 77.7% (17 mm), and 55.0% (10 mm) for 30 min and 85.7%, 78.1% and 53.9% for 15 min, respectively.

CRCs for ¹²⁴I exceeded those of the ¹⁸F reference for spheres ≥13 mm using SSS-TF, with a mean difference of 4.3 ± 3.1%, and a maximum deviation of 8.0% (37 mm).

Application of SSS-MLSS resulted in consistently lower CRC values compared to SSS-TF, with mean reductions of −12.5 ± 0.2% (30 min) and −10.6 ± 0.3% (15 min).

### Dependency of Quantitative Accuracy on Scan time, Scatter Correction and Object Geometry

Absolute quantification results are summarized in Table 2. For 30 min acquisitions using SSS-TF, representative RCs for ¹²⁴I were consistently lower than the corresponding values obtained for ¹⁸F across all sphere sizes (e.g. 77.1% vs. 88.7% for the 37 mm sphere and 47.5% vs. 60.0% for the 10 mm sphere), consistent with the larger positron range of ¹²⁴I. Scan time reduction to 15 min had no substantial impact on quantitative accuracy, with deviations below ±2% for both spheres and background (Table 2).

**Table 2.**
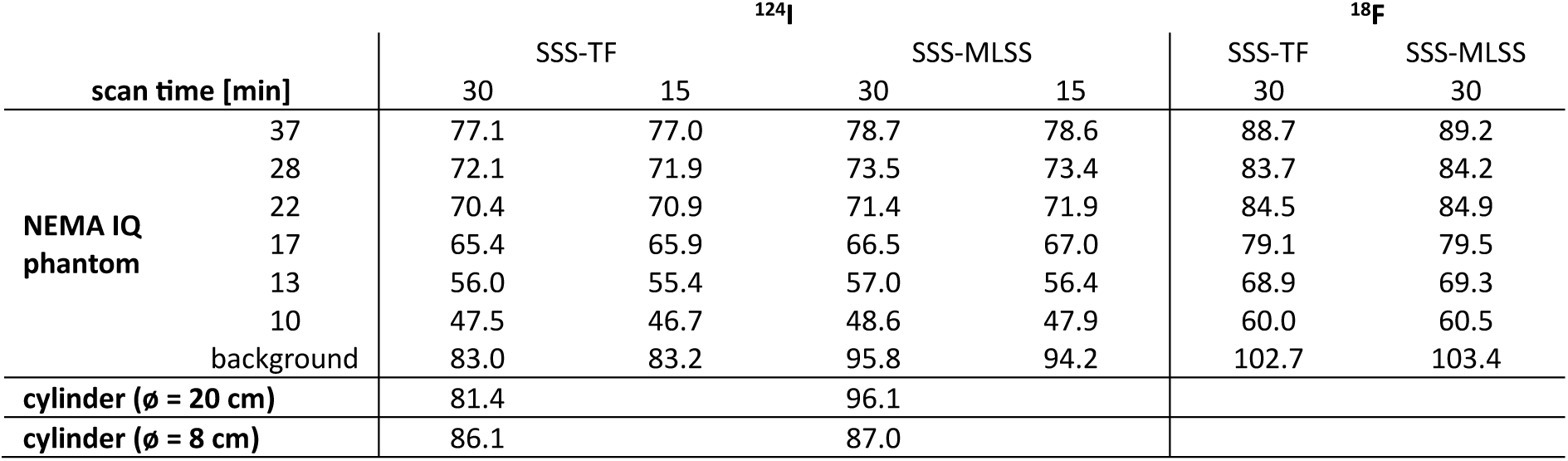
Quantification Accuracy in terms of recovery coefficients (RCs, [%]) for NEMA IQ phantom spheres and background compartment, as well as for the 20 cm and 8 cm diameter cylindrical phantoms. Results are shown for ^124^I (30 min and 15 min) using SSS-TF and SSS-MLSS, with ^18^F NEMA IQ phantom (SSS-TF and SSS-MLSS) included for comparison.

Consistent with the visual differences observed in the difference images (Figure 3C, F), absolute quantification of ¹²⁴I showed a systematic underestimation for both spheres and background when reconstructed using SSS-TF. Application of SSS-MLSS led to a marked increase in recovered activity in the background compartment of the NEMA IQ phantom, with RCs increasing from 83.0% (SSS-TF) to 95.8%, while only minor changes were observed in the spheres (+1.8 ± 0.3%). In contrast, only minor differences in sphere and background RC values between SSS-TF and SSS-MLSS were observed for ¹⁸F, suggesting that the pronounced effects seen for ¹²⁴I mainly originate from prompt gamma correction.

For the cylindrical phantoms no obvious visual differences between reconstruction methods were observed in the reconstructed images (Figure 5A, B, D, E). However, the corresponding difference images (Figure 5C, F) revealed again a systematic increase in activity for SSS-MLSS, with a spatial gradient showing higher deviations towards the center of the phantom.

**Figure 5.**
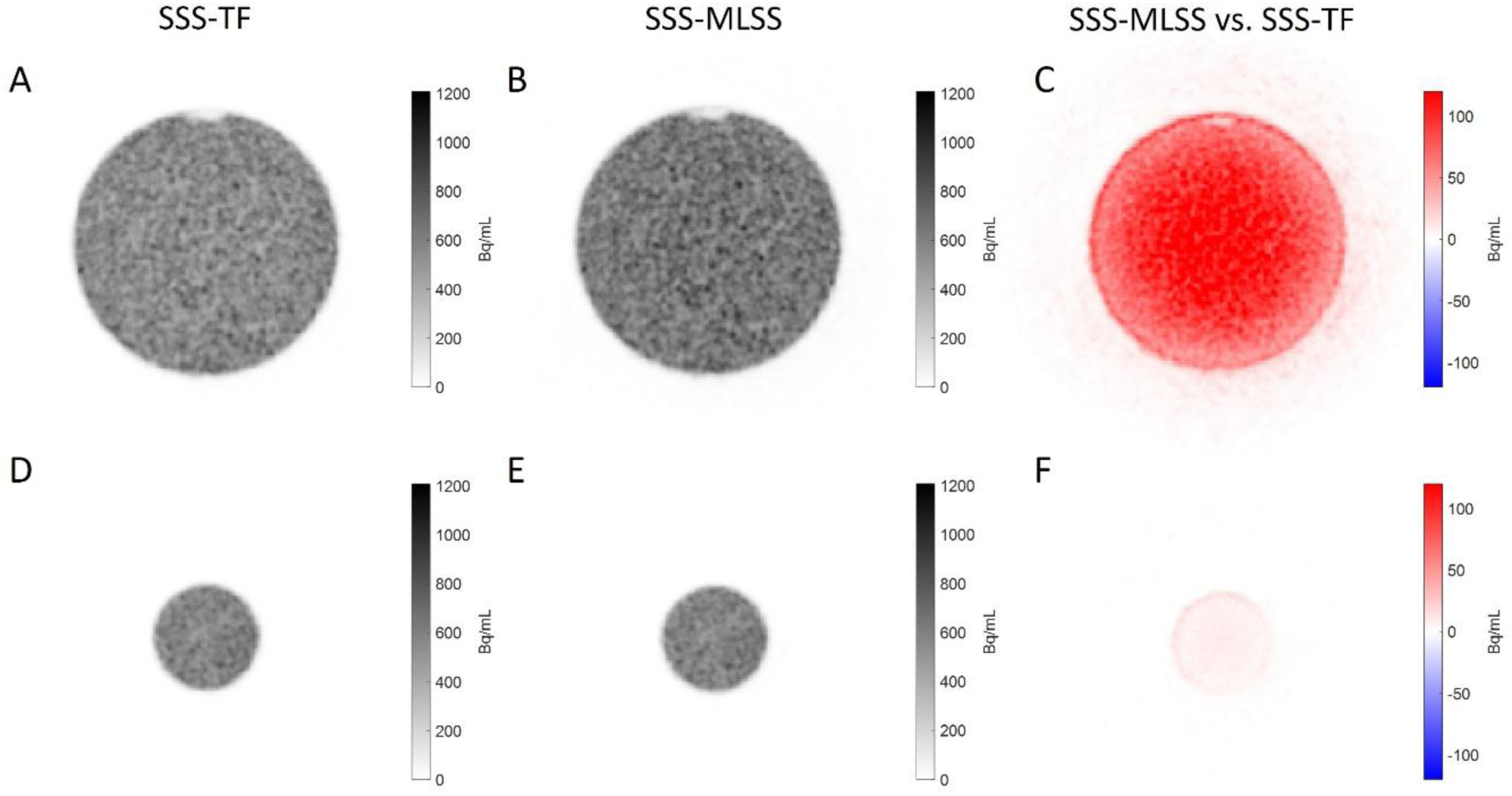
Representative transverse slices of the cylinder phantoms. (A–C) ¹²⁴I 20 cm cylinder reconstructed with SSS-TF (A), SSS-MLSS (B), and corresponding difference image (SSS-MLSS minus SSS-TF) (C). (D–F) ¹²⁴I 8 cm cylinder reconstructed with SSS-TF (D), SSS-MLSS (E), and corresponding difference image (F).

In line with the NEMA IQ results, size-dependent effects on quantification were observed in the cylindrical phantoms. In the larger 20 cm cylinder, SSS-TF underestimated the ^124^I activity with an RC of 81.4%, which was substantially less than using SSS-MLSS (RC: 96.1%). In contrast, for the smaller 8 cm cylinder, only minor differences were observed between SSS-TF (86.1%) and SSS-MLSS (87.0%), suggesting a reduced impact of prompt gamma correction in smaller objects.

### Patient Case

Lesion detectability remained preserved across all reconstruction methods and acquisition times, with all lesions clearly identifiable down to 5 min (Figure 6).

**Figure 6.**
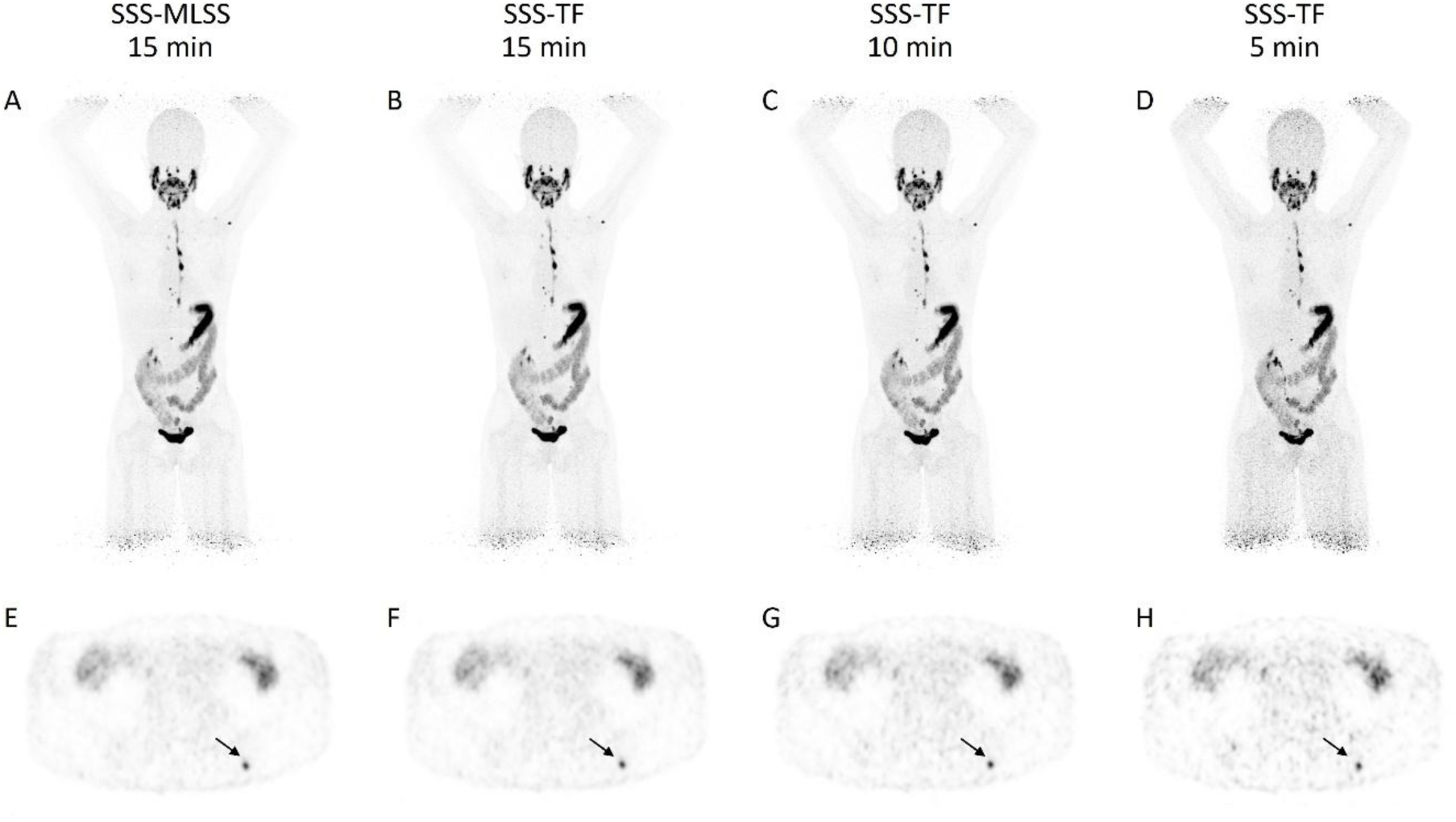
(A-D) Maximum intensity projection (MIP) PET images of a patient with metastatic differentiated thyroid cancer, acquired approximately 24 h after oral administration of 37 MBq ¹²⁴I on a LAFOV PET/CT system. Images were reconstructed using SSS-MLSS for 15 min (A) and SSS-TF for 15 min (B), 10 min (C), and 5 min (D). (E-H) Transverse slices of the os ilium metastasis (lesion #6, arrow) reconstructed using SSS-MLSS for 15 min (E; SUV_mean_ 2.76, SUV_max_ 3.81) and SSS-TF for 15 min (F; SUV_mean_ 2.78, SUV_max_ 3.84), 10 min (G; SUV_mean_ 3.27, SUV_max_ 4.92), and 5 min (H; SUV_mean_ 3.03, SUV_max_ 4.33).

Quantitative analysis demonstrated likewise a stable uptake in the lesions across the reconstruction methods, with comparable SUV_mean_ and SUV_max_ for SSS-MLSS (6.79 ± 4.73; 9.97 ± 7.31) and SSS-TF (6.76 ± 4.65; 9.97 ± 7.26) (Figure 7).

**Figure 7.**
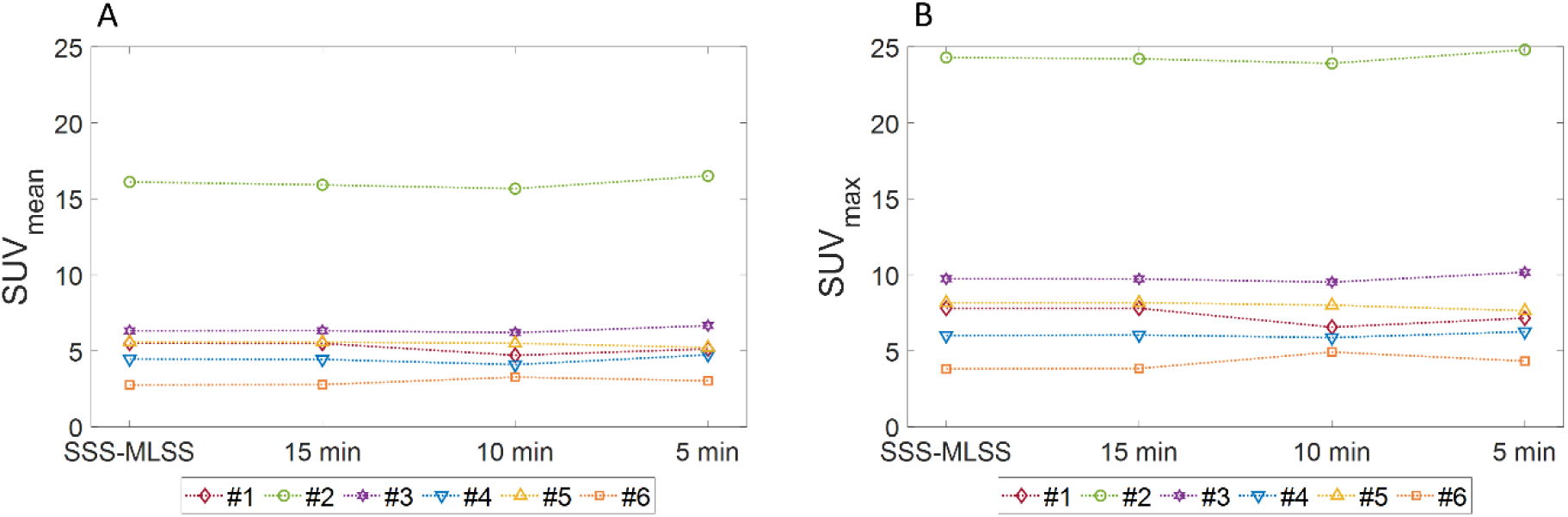
Quantitative analysis of ^124^I avid bone metastases of a patient with DTC. SUV_mean_ (A) and SUV_max_ (B) for individual lesions (#1-#6) reconstructed using SSS-MLSS (15 min) and SSS-TF (15 min, 10 min, 5 min).

Reduction of acquisition time to 10 min (SUV_mean_: 6.57 ± 4.57; SUV_max_: 9.80 ± 7.10) and 5 min (SUV_mean_: 6.88 ± 4.86; SUV_max_: 10.06 ± 7.47) resulted in only minor overall deviations. The largest variability was observed in the lesion of the os ilium (lesion #6) with deviations up to 17.6% in SUV_mean_ (3.27 vs. 2.78) and 28.1% in SUV_max_ (4.92 vs. 3.84) at 10 min (Figure 7).

## Discussion

This study systematically evaluated image quality and quantitative performance of low-count ¹²⁴I PET imaging on a LAFOV PET/CT system, with particular focus on potential sources of bias along the preparation, acquisition and data corrections in the reconstruction chain. Using dedicated phantom experiments under clinically realistic conditions, complemented by a clinical example of a DTC patient scan, the results demonstrate that LAFOV PET enables accurate ¹²⁴I imaging with preserved image quality and stable contrast recovery despite low activity concentrations, positron yield and reduced scan times. At the same time, moderate deviations in absolute quantification were observed, which could be attributed primarily to data corrections-related effects rather than limitations in count statistics or inaccuracies in activity calibration.

A central finding of this study is the robustness of image quality under low-count conditions representative for clinical ¹²⁴I imaging. Image noise using SSS-TF and 30 min acquisition time (CV 15.8%) closely matched the ∼15% benchmark recommended by EANM/EARL guidelines [30,31] and was comparable to matched low-count ¹⁸F acquisitions (CV 15.4%). This indicates that prompt gamma emissions and their correction do not substantially elevate image noise.

Even with reduced acquisition time (15 min), the contrast recovery remained largely stable (maximum deviation −2.1% for the 10 mm sphere), demonstrating preserved quantitative contrast despite substantially reduced count statistics.

This was further supported by the clinical dataset, where lesion detectability and visual contrast remained preserved even at reduced acquisition times down to 5 min, confirming the translational relevance of the phantom findings under realistic conditions.

These findings were consistent with previous work on LAFOV PET scanners, where strong count reduction (600 s to 15 s) resulted in only minor CRC deviations (±3% for spheres ≥13 mm) [17]. The presented CRC values for ^18^F (e.g. 85.6% and 80.5% for the 37 mm and 28 mm spheres, substantially lowered background of 0.1 kBq/ml) were in good agreement with previously reported ¹⁸F values (85.9% and 77.6%, 2.5 kBq/mL background) [17].

Notably, CRC values for ¹²⁴I exceeded those of ¹⁸F for spheres ≥13 mm, e.g. 92.5% vs. 85.6% for the 37 mm sphere. Similar findings have been reported by Preylowski et al. [19] for a conventional PET system, with CRC values of 74% for ^124^I and 65% for ^18^F for the 37 mm sphere. This apparent improvement does not reflect superior intrinsic performance of ¹²⁴I imaging. It rather can be attributed to differences in absolute quantification between spheres and background.

To this end, a systematic underestimation of activity concentration was observed for ¹²⁴I using SSS-TF, affecting both spheres and background, but more pronounced in the background (83.0% for ^124^I vs. 102.7% for ¹⁸F), while the sphere recovery showed only smaller deviations (72.1% vs. 83.7% for the 28 mm sphere).

This is consistent with previously reported background underestimation of ∼20% for ¹²⁴I on a Biograph mCT system [20] and explains the elevated CRC values observed for ¹²⁴I, highlighting the limitations of ratio-based metrics in the presence of non-uniform bias.

The application of the alternative scatter correction method (SSS-MLSS) substantially improved the background recovery, increasing RC from 83.0% (SSS-TF) to 95.8%, while largely preserving sphere recovery (e.g. 78.7% vs. 77.1% for the 37 mm sphere). Consequently, CRC values were reduced compared to SSS-TF. Nevertheless, CRC values for ¹²⁴I remained consistently lower than those for ¹⁸F across all sphere sizes (e.g. 17 mm sphere: 67.8% for ¹²⁴I vs. 75.6% for ¹⁸F; 10 mm sphere: 48.2% vs. 56.2%), which is consistent with the larger positron range of ¹²⁴I compared with ¹⁸F (mean positron range in water: ∼2.8-4.4 mm for ¹²⁴I vs. ∼0.6 mm for ¹⁸F [22]) . Similar findings have previously been reported for SAFOV PET systems [19,32].

The improved quantification using SSS-MLSS is likely related to its global scaling approach, which utilizes the full emission data instead of relying on sinogram tail fitting. In contrast, SSS-TF may be more susceptible to inaccuracies under low-count conditions or when tail regions are limited or biased, for example in large objects with high FOV occupancy [33]. While previous studies using ¹⁸F reported only minor differences between both methods [34,35], the larger discrepancy observed here for ¹²⁴I likely reflects the influence of prompt gamma emissions, which contribute to elevated tail activity and may bias tail fitting.

Importantly, the observed differences between SSS-TF and SSS-MLSS for ¹²⁴I should not be interpreted as reflecting scatter correction alone. In the employed reconstruction framework, prompt gamma correction is intrinsically coupled to the scatter scaling procedure. The minimal differences observed between SSS-TF and SSS-MLSS for ¹⁸F in this study indicate that the scatter correction methodology itself contributes only marginally to the observed discrepancies. In contrast, the substantially larger differences observed for ¹²⁴I suggest that prompt gamma correction represents the dominant source of reconstruction-dependent quantification bias.

This interpretation is supported by the cylindrical phantom experiments in this study, where recovery with SSS-TF decreased with increasing object size (8 cm: 86.1% vs. 20 cm: 81.4%), consistent with increased scatter fraction and prompt gamma contribution in larger objects. In contrast, SSS-MLSS restored recovery in the larger, patient-sized phantom to 96.1%, indicating improved robustness with respect to object size and scatter conditions.

From a clinical perspective, this suggests that quantification bias in ¹²⁴I PET is object-dependent and primarily affects low-activity background regions. This is consistent with the clinical dataset, where lesion quantification remained largely unaffected by the choice of scatter correction, with comparable SUV_mean_ and SUV_max_ values for SSS-MLSS (6.79 ± 4.73 and 9.97 ± 7.31) and SSS-TF (6.76 ± 4.65 and 9.97 ± 7.26). These findings suggest that reconstruction-dependent bias observed in the phantom background may have only limited impact on focal lesion quantification in clinical ¹²⁴I imaging. Of note, these observations are based on a single clinical case and should therefore be interpreted with caution.

However, improved absolute quantification with SSS-MLSS was accompanied by increased residual activity in cold regions, reflected by an increase in lung residual error from 1.9% to 7.0% for ^124^I and from 4.1% to 6.3% for ^18^F. Similar effects have been reported for ¹⁸F imaging on the same system [34,35] indicating a trade-off between improved absolute quantification and reduced accuracy in cold regions.

Potential sources of error along the calibration chain were also evaluated. Although accurate dose calibrator calibration for ¹²⁴I is known to be challenging due to its complex emission spectrum and geometry dependence [25,36,37], measurements in this study demonstrated stable performance across a wide activity range, with low inter-device variability (1.6–2.3%) and good agreement down to ∼1 MBq. These findings indicate that, under controlled conditions, calibration-related uncertainties contribute only marginally to the observed deviations.

An alternative, more easily implementable approach [38] adjusts dose calibrator settings based on PET-derived activity measurements. While this may improve internal consistency, it inherently propagates reconstruction-related biases into absolute quantification, such as for SSS-TF in this study.

Overall, the presented results indicated that, with appropriate calibration procedures the primary sources of bias in ^124^I PET quantification arise from data correction methods in the image reconstruction, particularly prompt gamma and scatter modeling in the presence of prompt gamma contamination.

Beyond these effects, radionuclide-specific factors such as the extended positron range of ¹²⁴I further impact the quantitative accuracy by reducing spatial resolution and contrast. Previous work has shown that spatially variant positron range correction can improve contrast recovery by 4–24% [39], suggesting additional potential for optimization.

Taken together, these results demonstrate that LAFOV PET enables accurate low-count ¹²⁴I imaging with preserved image quality, while quantitative accuracy is primarily governed by prompt gamma and scatter correction methodology and object-dependent effects rather than calibration uncertainties.

These findings further suggest that acquisition protocols with reduced acquisition time may be feasible without compromising lesion detectability or quantitative assessment, which is particularly relevant for patient comfort and clinical workflow.

## Conclusion

This study demonstrates that LAFOV PET enables robust ¹²⁴I imaging under low-count conditions typical of clinical imaging, with preserved image quality and contrast even at reduced acquisition times ≤15 min.

Quantitative accuracy, however, was primarily influenced by prompt gamma and scatter correction methodology, with conventional approaches leading to systematic underestimation, particularly in the background. The use of SSS-MLSS improved absolute quantification and reduced bias, although at the expense of increased residual activity in low-count regions.

In contrast, dose calibrator performance and cross-calibration were stable and contributed only marginally to the observed deviations when appropriate calibration procedures were applied.

Overall, these findings indicate that accurate ¹²⁴I quantification on LAFOV systems is feasible but requires careful selection of reconstruction methods and calibration.

## Data Availability

The datasets used and/or analyzed during the current study are available from the corresponding author on reasonable request.

## Declarations

### Ethics approval and consent to participate

The local Institutional Review Board approved the study (773/2022BO2). The study was performed in accordance with the Declaration of Helsinki.

### Consent to participate

Written informed consent was obtained from the participant included in the study.

### Consent to publish

The participant provided written informed consent for publication.

### Competing Interests

JC is a full-time employee of Siemens Medical Solutions USA, Inc. FS and ClF received a research grant from Siemens Healthineers. IR received research funding not related to this study through an agreement between Siemens Healthineers and the Medical University of Vienna. There are no other conflicts of interest to report.

### Funding

This research was supported by the medical faculty of Eberhard Karls University Tübingen and the Ministry for Science, Research and the Arts Baden–Württemberg. The total-body PET/CT scanner was funded by the Deutsche Forschungsgemeinschaft (DFG, German Research Foundation)—INST 37/1145-1 FUGG. Furthermore, this research was funded by Deutsche Forschungsgemeinschaft (DFG, German Research Foundation, Germanýs Excellence Strategy-EXC-2180-390900677).

### Authors’ contributions

Conceptualization: EK, WJ, IR, FS; data curation: EK, PL, FS; formal analysis: EK, WJ, JS; funding acquisition: ClF, FS; investigation: EK, WJ, PL, IR, ClF, FS; methodology: EK, WJ, PL, JC, JS, FS; project administration: ClF, FS; resources: ClF, FS; software: EK, FS; supervision: FS; validation: EK, FS; visualization: EK; writing—original draft: EK, FS; writing—review and editing: EK, WJ, PL, JC, JS, IR, ClF, FS. All authors read and approved the final manuscript.

## Notes

### Author Declarations

The Institutional Review Board of the Medical Faculty of the Eberhard Karls University and the University hospital Tuebingen approved the study (773/2022BO2). The study was performed in accordance with the Declaration of Helsinki.

